# Synthesizing evidence regarding artificial intelligence generated radiological reports based on medical images: a scoping review protocol

**DOI:** 10.1101/2025.03.17.25324103

**Authors:** Weibo Feng, Anthony Yazdani, Alban Bornet, Alexandra Platon, Douglas Teodoro

## Abstract

**Introduction:** Considering numerous radiological images and the heavy workload of writing corresponding reports in clinical work, it is significant to leverage artificial intelligence (AI) to facilitate this process and reduce the burden of radiologists. In the past few years, particularly with the advent of vision language models, some works explored generating radiological reports directly from images. However, despite some efforts demonstrated in previous studies, limitations in AI-generated radiological reports persist. Current research mainly focuses on detecting abnormalities, rather than generating textual reports from medical images. The evidence for AI application in radiological report writing has not been synthesized. This scoping review aims to map the current literature on the engagement of AI-generating radiological reports based on images.

**Methods and analysis:** Following a well-established scoping review methodology, five stages are provided: i) determining the research question, ii) searching strategy, iii) inclusion/exclusion criteria, iv) data extraction, and v) results analysis. Four databases will be applied to search peer-reviewed literature from January 2016 to February 2025. A two-stage screening process will be conducted by two independent reviewers to determine the eligibility of articles, and only those regarding AI-generated radiological reports will be included. All data from eligible articles will be extracted and analyzed using narrative and descriptive analyses, presenting in a standard form.

**Ethic and dissemination:** Ethic approval is no required in this scoping review. Experts from Hospital of University of Geneva will be consulted to provide professional insight and feedback regarding the study findings and help with dissemination activities in peer-reviewed publications or academic presentations

## Introduction

Medical images, especially radiological images, such as computed tomography (CT), magnetic resonance imaging (MRI), X-ray, etc., play an imperative role in clinical screening, diagnosis, and treatment. Generally, medical images should be presented pair with corresponding textual reports, explaining some basic information, findings or conclusions to both patients and doctors. There are several sections in a structured radiological report, including *basic information, indication, description* and *conclusion. Basic information* and *indication* part should be fulfilled automatically in system, while *description* and *conclusion* sections consume most of radiologists’ time. In clinical scenario, radiologists routinely view and analyze various images, then describe the findings or abnormalities in *description* section and make a summary in *conclusion* section for each case, which is a very tedious, time consuming and error-prone task. As a rapid growth of radiological images in recent years, how to assist radiologists in writing reports and reduce their working burden becomes an important and challenging task, especially in situations where number of radiologists is low and medical resources are limited.

In the last decade, artificial intelligence (AI), one of the most disruptive technologies, has been widely applied in medical images and some promising achievements have been made^[1]^. Deep learning, such as convolutional neural network (CNN) and recurrent neural network (RNN) has been proved effective in processing large-scale multimodal medical images and detecting abnormalities^[2-5]^. Natural language processing (NLP), especially vision language models (VLM), can be used to combine medical images and text, generating medical image reports^[6-7]^. With the goal to support radiologists in reducing workload and diagnostic errors, some tasks of automagical radiology report generation from medical images are proposed with NLP^[8-9]^.

However, irrespective of some impressive findings in current research, it is still challenging to leverage AI to generate clinically readable and interpretable reports in long complex structures based on radiological images. Most existing AI models perform better in identifying abnormalities or generating short descriptions of images^[10-14]^, which provides very limited help to radiologists in real clinical practice. On the other hand, patients also need some brief, interpreting, and easy-understanding reports about radiological images as well.

Several attempts to alleviate workload and facilitate both patients and radiologists by AI generating reports directly from radiological images are emerging^[8-9,15-17]^, but the extent and nature of this evidence remain unclear currently. Hence, some efforts have been made to provide preliminary summaries in recent reviews about AI application in radiology. However, the majority of previous reviews either focused on one specific subspecialty including nuclear medicine, musculoskeletal imaging, etc.^[18-19]^, or based on one specific AI model like Chat Generative Pre-Trained Transformer (ChatGPT), Bidirectional Encoder Representations from Transformers (BERT), etc.^[20-21]^, or restrict to one specific type of radiological image such as X-ray[22]. A few other reviews also summarized recent advancements in VLM for medical report generating, focusing on publicly available models, architecture, public dataset and evaluation metric for VLM^[23]^, while current application scope in radiology is not fully synthesized. Hence, more comprehensive overview about the application scope or coverage of AI-generating reports in radiology and how radiologists and patients can benefit from current AI technologies remains inadequate for now.

To address this knowledge gap, we aim to map the available literature from January 2016 to February 2025 regarding AI-generated radiological reports based on medical images in this scoping review. In contrast to previous related surveys, this review aims to provide a comprehensive update regarding how AI is integrated with radiological images to facilitate medical report generation with detailed subspecialties, context, use-cases, algorithms, image types and performances. In doing so, we hope this scoping review will inform future radiological AI development and identify areas of future research.

## Methods and analysis

In this scoping review, we will follow Preferred Reporting Items for Systematic Reviews and Meta-Analysis extension for Scoping Reviews (PRISMA-SCR) guidelines^[24]^ and make use of literature across both scientific journal and conference peer-reviewed literature from January 2016 to February 2025 in Pubmed, Embase, Web of Science and Google Scholar. There five stages in this study: (1) determining the research question, (2) search strategy, (3) inclusion/exclusion criteria, (4) data extraction, (5) analysis and presentation of the results. Ethic review board approval is not required in this study.

### Research question

Group members of this study had previously convened experts in a consensus meeting to identify high-priority research questions specific to generating AI radiological reports directly from medical images. After rigorous discussion, this scoping review is designed to answer the following research questions:

1. In which subspecialties and conditions AI is being used for generating radiological reports from medical images?
2. What are the use-cases (e.g., structure vs. free-text) and context (e.g., clinical-vs. patient-oriented report) in which AI models are being used for generating radiological reports from medical images?
3. What are the AI algorithms used for generating radiological reports based on medical images?
4. What kind of medical images, such as CT, MRI or X-ray, are used as data sources for AI model training?
5. What are the performances and limitations of AI models for generating radiological reports from images?

### Search strategy

After careful consideration, 4 databases were selected in scoping review: Pubmed, Embase, Web of Science and Google Scholar. The query definitions for searching keywords are shown in Tab.1.

**Tab.1.** Keywords and query definitions for search

|  |  |
| --- | --- |
| Group 1-Methodology related keywords | “artificial intelligence” OR “deep learning” OR “neural network” OR “natural language processing” OR “language model” OR “vision model” OR “transformers” OR “diffusion model” OR “generative model” OR “CNN” OR “RNN” OR “LSTM” OR “ViT” OR “LLM” OR “VLM” OR “NLP” OR “report* generation model” OR “report* generation algorithm” OR “text decoder” OR “text encoder” OR “image encoder” |
| Group 2-Medicine related keywords | “Computed tomography” OR “Magnetic Resonance Imaging” OR “Radiography” OR “X-ray” OR “Thoracic” OR “Mammography” OR “Diagnostic imaging” OR “Neuroimaging” OR “Angiography” OR “Musculoskeletal radiology” OR “Chest imaging” OR “Oncologic imaging” OR “Abdominal imaging” OR “Breast imaging” |
| Group 3 -Task related keywords | “generating” OR “generation” OR “write” OR “writing” OR “creation” OR “creating” OR “formatting” AND “report” |
| Final results | (Group 1) AND (Group 2) AND (Group 3) |

All literature searches will be conducted by an information scientists in our team and a panel of radiologists will be consulted to ensure all relevant citations are obtained. All literatures included in this review will be amalgamated using EndNote to ensure there are no duplicates in database.

### Inclusion criteria

#### Concept

Radiological report is defined as a textual file describing findings or abnormalities in corresponding radiological images and making some preliminary summaries. Structured report is defined as a report writing contents with structured sentences in a set template or routine pattern. Free-text report is defined as a report writing in free choices of sentences, paragraphs without set structure or template.

#### Medical purpose

Generating AI radiological reports based on corresponding medical images for radiologists or patients.

#### Methodology

Natural language models and other related AI technologies focused on generating radiological textual reports.

#### Dataset

Various kinds of radiological images used in common clinical scenario, such as X-ray, CT, MRI.

#### Article type

Basic research and peer-reviewed articles published in journals or conferences. Language: English.

#### Period

01.01.2016-28.02.2025

### Exclusion criteria

No structured or free-text radiological reports are generated using AI models.

AI radiological reports are generated based on other textual datasets instead of images. Generating other medical reports, such as pathological reports, based on images.

### Data extraction

After the search of eligible literatures in all databases, a knowledge synthesis management platform, Covidence, will be used for screening and deduplication. Title and abstract screening will be conducted in duplicate on all involved articles and 2 of our reviewers will independently categories each article as ‘Yes’, ‘No’, or ‘Maybe’ label according to our inclusion and exclusion criteria. Articles with ‘Yes’ or ‘Maybe’ will be included for further full text screening and extraction. If any discrepancies raise, another senior reviewer in our team will be consulted. For those included for full-text screening, 2 research team members will independently review and categories the articles as either ‘Yes’ or ‘No’. Any uncertainties will be discussed by the whole team members. After full-text reviewing, data extraction will be completed by specified team members for all included articles following the extraction plan shown in Tab.2.

**Tab.2.**
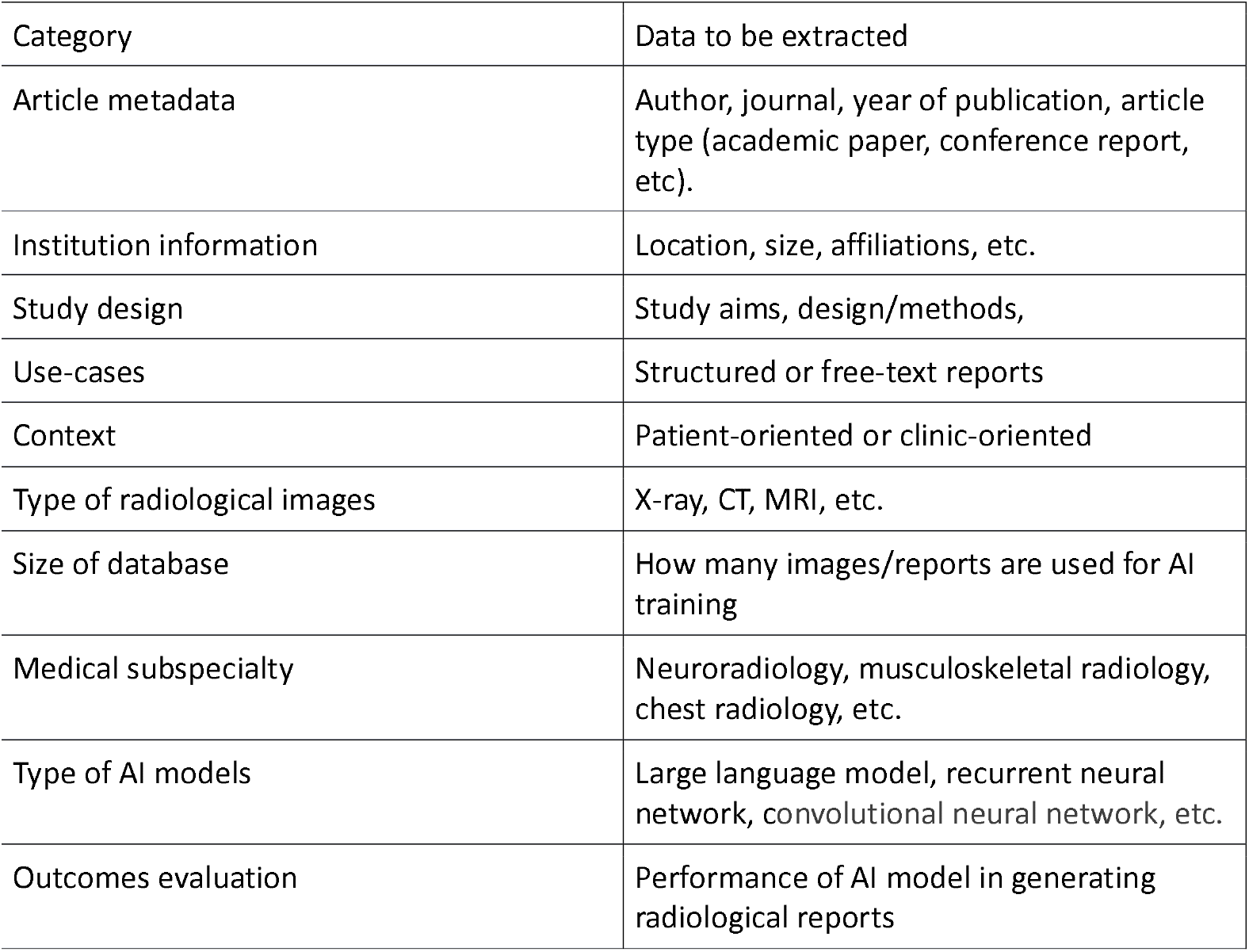
Preliminary data extraction plan

### Analysis and presentation of results

In order to provide a clear overview of the extent and results, we will employ a modified PRISMA-SR, a basic numerical summary of type and distribution of articles included, as well as a visual chart presenting the key data extraction categories. According to the searching and screening results, we will apply a framework that best summary and present the results.

## Data Availability

All data produced in the present study are available upon reasonable request to the authors

## Knowledge translation

In this scoping review, experts of radiology and AI algorithm in University of Geneva focused on AI training and writing medical reports will be consulted to help us in dissemination of results. We aim to provide insight and summary of current study findings, engaging in the development of future research. Traditional dissemination activities, including peer-reviewed publications, academic presentations at conferences are planned.

## Ethic and patient involvement

No patients or public are involved in this scoping review and ethics approval is not applicable.

## Reference

1. Interagency Working Group on Medical Imaging 1Committee on Science (2017). National Science and Technology Council, Roadmap for Medical Imaging Research and Development, Washington, D.C., USA, pp. 1–19.

2. L. Ebner, M. Tall, K.R. Choudhury, et al. (2017). Variations in the functional visual field for detection of lung nodules on chest computed tomography: impact of nodule size, distance, and local lung complexity, Med. Phys. 44, 3483–3490.

3. W. Sun, B. Zheng, W. Qian (2017). Automatic feature learning using multichannel ROI based on deep structured algorithms for computerized lung cancer diagnosis, Comput. Biol. Med. 89, 530.

4. W. Sun, T.B. Tseng, J. Zhang, et al. (2017). Enhancing deep convolutional neural network scheme for breast cancer diagnosis with unlabeled data, Comput. Med. Imaging. Graph. 57, 4–9.

5. A. Masood, B. Sheng, P. Li, et al. (2018). Computer-assisted decision support system in pulmonary cancer detection and stage classification on CT images, J. Biomed. Inform. 79, 117–128.

6. Kisilev P, Walach E, Barkan E, Ophir B, Alpert S, Hashoul SY (2015) From medical image to automatic medical report generation. IBM J Res Dev 59(2/3):2:1–2:7

7. Li, B., Zhang, Y., Guo, D., Zhang, R., Li, F., Zhang, H., Zhang, K., Li, Y., Liu, Z., and Li, C. (2024). Llava-onevision: Easy visual task transfer. arXiv preprint arXiv:2408.03326.

8. Kisilev P, Sason E, Barkan E, Hashoul S (2016). Medical image description using multi-task-loss CNN. In: Carneiro G, Mateus D, Peter L, Bradley A, Tavares JMRS, Belagiannis V, Papa JP, Nascimento JC, Loog M, Lu Z, Cardoso JS, Cornebise J (eds) Deep learning and data labeling for medical applications. Springer, Berlin, pp 121–129

9. Jing B, Xie P, Xing E (2018). On the automatic generation of medical imaging reports. In: Proceedings of the 56th annual meeting of the association for computational linguistics (volume 1: long Papers). Association for Computational Linguistics, pp 2577–2586

10. P. Kisilev, E. Walach, E. Barkan, et al. (2015). From medical image to automatic medical report generation, IBM J. Res. Dev. 59, 2:1–2:7.

11. H.C. Shin, K. Roberts, L. Lu, et al. (2016). Learning to read chest X-rays: recurrent neural cascade model for automated image annotation, in IEEE Conference on Computer Vision and Pattern Recognition, IEEE Computer Society, Las Vegas, NV, USA, pp.2497–2506.

12. Singhal, K., Azizi, S., Tu, T., Mahdavi, S. S., Wei, J., Chung, H. W., Scales, N., Tanwani, A., Cole-Lewis, H., Pfohl, S., et al. (2023). Large language models encode clinical knowledge. Nature, 620(7972):172–180.

13. Li, C., Wong, C., Zhang, S., Usuyama, N., Liu, H., Yang, J., Naumann, T., Poon, H., and Gao, J. (2023a). Llava-med: Training a large language-and-vision assistant for biomedicine in one day. arXiv preprint arXiv:2306.00890.

14. Saab, K., Tu, T., Weng, W.-H., Tanno, R., Stutz, D., Wulczyn, E., Zhang, F., Strother, T., Park, C., Vedadi, E., et al. (2024). Capabilities of gemini models in medicine. arXiv preprint arXiv:2404.18416.

15. Nooralahzadeh, F., Perez Gonzalez, N., Frauenfelder, T., Fujimoto, K., and Krauthammer, M. (2021). Progressive transformer-based generation of radiology reports. In Moens, M.-F., Huang, X., Specia, L., and Yih, S.W.-t., editors, Findings of the Association for Computational Linguistics: EMNLP 2021, pages 2824–2832, Punta Cana, Dominican Republic. Association for Computational Linguistics.

16. Alfarghaly, O., Khaled, R., Elkorany, A., Helal, M., and Fahmy, A. (2021). Automated radiology report generation using conditioned transformers. Informatics in Medicine Unlocked, 24:100557.

17. Tanida, T., Müller, P., Kaissis, G., and Rueckert, D. (2023). Interactive and explainable region-guided radiology report generation. In Proceedings of the IEEE/CVF Conference on Computer Vision and Pattern Recognition, pages 7433–7442

18. Gorelik, N., & Gyftopoulos, S. (2021). Applications of Artificial Intelligence in Musculoskeletal Imaging: From the Request to the Report. Canadian Association of Radiologists journal = Journal l’Association canadienne des radiologistes, 72(1), 45–59

19. Hirata, K., Matsui, Y., Yamada, A., Fujioka, T., Yanagawa, M., Nakaura, T., Ito, R., Ueda, D., Fujita, S., Tatsugami, F., Fushimi, Y., Tsuboyama, T., Kamagata, K., Nozaki, T., Fujima, N., Kawamura, M., & Naganawa, S. (2024). Generative AI and large language models in nuclear medicine: current status and future prospects. Annals of nuclear medicine, 38(11), 853–864.

20. Meşe, İ., Altintaş Taşliçay, C., Kuzan, B. N., Kuzan, T. Y., & Sivrioğlu, A. K. (2024). Educating the next generation of radiologists: a comparative report of ChatGPT and e-learning resources. Diagnostic and interventional radiology (Ankara, Turkey), 30(3), 163–174.

21. Gorenstein, L., Konen, E., Green, M., & Klang, E. (2024). Bidirectional Encoder Representations from Transformers in Radiology: A Systematic Review of Natural Language Processing Applications. Journal of the American College of Radiology : JACR, 21(6), 914–941.

22. Ouis, M. Y., & A Akhloufi, M. (2024). Deep learning for report generation on chest X-ray images. Computerized medical imaging and graphics : the official journal of the Computerized Medical Imaging Society, 111, 102320.

23. Hartsock, I., & Rasool, G. (2024). Vision-language models for medical report generation and visual question answering: a review. Frontiers in artificial intelligence, 7, 1430984.

24. Tricco AC, Lillie E, Zarin W, et al. (2018). PRISMA extension for scoping reviews (PRISMA-SCR): checklist and explanation. Ann Intern Med; 169:467–73

